# Favourable discharge after cardiac arrest across APACHE IVa-predicted hospital mortality: a multicentre observational study

**DOI:** 10.64898/2026.09.13.26362943

**Authors:** Alon Gorenshtein, Amir Srour, Yosef Adiniaev, Yiftach Barash, Eyal Klang, Oved Daniel

**Affiliations:** Department of Neurology, Beth Israel Deaconess Medical Center, Harvard Medical School, Boston, MA, USA; BRIDGE GenAI Lab, Beth Israel Deaconess Medical Center, Boston, MA, USA; Department of Radiology, Beth Israel Deaconess Medical Center, Harvard Medical School, Boston, MA, USA; Neurology Division, Tel Aviv Sourasky University Medical Center, Tel Aviv, Israel

**Keywords:** cardiac arrest, neuroprognostication, functional outcome, illness severity scores, withdrawal of life-sustaining therapy

## Abstract

**Aim:** General ICU severity scores predict hospital mortality across heterogeneous populations, not recovery within one diagnosis. We described favourable discharge across the range of APACHE IVa-predicted hospital mortality after cardiac arrest, and tested whether any bedside subgroup fell below a pre-specified benchmark.

**Methods:** Observational cohort of 3,641 first ICU admissions in 161 eICU hospitals with a cardiac-arrest diagnosis documented within 24 hours. The exposure was APACHE IVa-predicted hospital mortality; the outcome was favourable discharge (home or rehabilitation), a functional-outcome proxy. Fourteen bedside subgroups were pre-specified; the benchmark required an upper 95% bound below 5%.

**Results:** Favourable discharge occurred in 1,039 (28.5%), falling from 78.9% in the lowest predicted-mortality decile to 4.1% in the highest (trend z, -27.2; P < 0.001). Among 2,139 patients with predicted mortality at or above 50%, 301 (14.1%; 95% CI, 12.6 to 15.6) had a favourable discharge; among 281 at or above 90%, 9 (3.2%) did. No subgroup had an upper 95% bound below 5%; the lowest was 8 of 264 (3.0%; 95% CI, 1.3 to 5.9). At a more permissive 10% benchmark, two qualified. Among 1,961 not obeying commands, 263 (13.4%) had a favourable discharge. Discrimination did not differ detectably between unfavourable functional outcome (AUROC 0.789) and in-hospital death (0.774; P = 0.078).

**Conclusion:** Favourable discharge declined as predicted mortality rose but remained observable at the highest risk levels, and no pre-specified subgroup was bounded below 5%. A general ICU mortality estimate is not on its own evidence that a favourable outcome is absent.

## 1. Introduction

Acute Physiology and Chronic Health Evaluation (APACHE) IVa and comparable general severity scores are routinely calculated for eligible intensive care admissions to estimate hospital mortality. APACHE IVa was developed and calibrated to predict in-hospital death across the general intensive care population, and it summarises illness severity across heterogeneous populations rather than prognosis within any one diagnosis.^1^

After cardiac arrest that distinction carries unusual weight. Guidelines recommend multimodal prognostication and advise considering formal assessment of poor neurological outcome in patients not awake and obeying commands at 72 hours or later after return of spontaneous circulation, once major confounders have been considered.^2,3^ A general severity score is derived from the first intensive care day and is designed neither for prognosis within a specific diagnosis nor for functional recovery rather than survival. Decisions taken early can foreclose the outcome they anticipate, since most in-hospital deaths after cardiac arrest follow withdrawal of life-sustaining therapy.^4,5^

APACHE II discriminated mortality and neurological outcome in a prospective post-arrest cohort,^6^ and a later prospective study compared APACHE II and SAPS II against arrest-specific scores.^7^ Those studies asked how well such scores rank outcome in cohorts of a few hundred patients. Purpose-built post-arrest scores use variables specific to arrest and neurological injury,^8,9^ but require deliberate collection. What has not been described at scale is the absolute probability of a favourable discharge across the full range of a general severity estimate, or whether any pre-specified bedside subgroup bounds that probability below a low benchmark. We conducted a cohort study across 161 hospitals to describe favourable discharge across the full range of APACHE IVa-predicted hospital mortality after cardiac arrest, and to test pre-specified bedside subgroups against a low-recovery benchmark.

## 2. Methods

### 2.1 Study Design and Setting

This was a retrospective observational cohort study of three public intensive care databases, reported according to the STROBE statement and its RECORD extension for routinely collected health data.^10,11^ No intervention was assigned and no causal effect was estimated. The analysis plan, including the primary outcome, the high-risk threshold, the four primary tests, the 14 subgroups and the low-recovery benchmark, was committed to the authors’ version-controlled repository before the subgroup results were generated (Supplementary S1.14).

### 2.2 Cohort Identification

The primary eICU cohort comprised first intensive care unit stays with a diagnosis explicitly naming cardiac arrest, first documented no more than 1,440 minutes after admission, with a recorded hospital discharge disposition and an APACHE IVa predicted hospital mortality between 0 and 1 (Supplementary S1.1 and S1.3). A broader screening cohort that also admitted anoxic and hypoxic-ischaemic diagnoses without a named arrest, and tighter diagnosis-timing windows of 360 and 60 minutes, were sensitivity analyses. MIMIC-IV admissions required an International Classification of Diseases code for cardiac arrest (ICD-9 4275, ICD-10 I46). No age, sex or admission-year restriction was applied.

### 2.3 Data Sources and Variables

The primary cohort came from the eICU Collaborative Research Database v2.0, a multicentre intensive care database.^12^ Replication used MIMIC-IV v3.1, an independent single-health-system database.^13^ An outcome anchor used the International Cardiac Arrest Research Consortium (I-CARE) database, in which Cerebral Performance Category was recorded prospectively.^14,15^ The public eICU release is a stratified sample with unpublished per-hospital sampling fractions, so we report proportions and associations only, and no rate or per-hospital quantity (Supplementary S1.4).

Bedside covariates were age, sex, Glasgow Coma Scale eye, motor and verbal response from the APACHE physiology record, intubation and mechanical ventilation status, and arterial pH. In MIMIC-IV, motor response was the first value recorded for each admission (item 223901).

### 2.4 Exposure and Outcomes

The exposure was APACHE IVa predicted hospital mortality as recorded, without rescaling or refitting. The pre-specified high-risk stratum was predicted mortality at or above 0.50, chosen a priori as an intuitive boundary at which predicted mortality equals predicted survival rather than as a clinical treatment threshold; results are also reported at 0.70 and 0.90.

The primary outcome was favourable discharge, a proxy for functional outcome, defined as discharge home or to rehabilitation. All other dispositions, including death in hospital, were unfavourable (Supplementary S1.5). Discharge disposition after cardiac arrest is associated with Cerebral Performance Category and modified Rankin Scale at discharge and is used as a functional-outcome proxy where formal scales are not recorded.^16^ Discharge home only and survival to hospital discharge were carried through every analysis as sensitivity outcomes.

### 2.5 Statistical Analysis

Proportions carry exact (Clopper-Pearson) 95% confidence intervals,^17^ and every primary proportion also carries a hospital-clustered interval from a nonparametric bootstrap resampling hospitals over 5,000 replicates. Trend across deciles of predicted mortality, and across four pre-specified age bands within the high-risk stratum, was tested with the Cochran-Armitage test. Favourable discharge in the high-risk stratum was compared against 5% with a one-sided exact binomial test.

We pre-specified 14 bedside subgroups from age, Glasgow Coma Scale motor response, mechanical ventilation, intubation and predicted mortality, analysing each subgroup with at least 20 patients. A subgroup met the low-recovery benchmark only when the upper limit of its 95% confidence interval fell below 5%, on both the exact and the hospital-clustered interval; the classification rested on the bound, not the point estimate. The 5% value is an analytic benchmark, not a clinical threshold for withdrawal of life-sustaining therapy, and classification is also reported at a more permissive 10%. Because the rule tests an upper bound, failure at 5% entails failure at any stricter benchmark (Supplementary S1.6).

Within the high-risk stratum, the adjusted association of age (per 10 years), sex, motor response and mechanical ventilation with favourable discharge was estimated by generalised estimating equations with hospital as the cluster (Supplementary S1.8); these coefficients are exploratory and their P values unadjusted. Discrimination for unfavourable functional outcome was compared with discrimination for in-hospital death, the endpoint APACHE IVa was built to predict, by a paired bootstrap over 2,000 replicates resampling hospitals.

Four primary tests were named in advance and adjusted together with the Benjamini-Hochberg procedure.^18^ The subgroup search is an interval-based classification and carries no significance test. Secondary sensitivity analyses sat outside the multiplicity family and are specified in Supplementary S1.11; the timing, admission-source, admission-diagnosis and survivor-conditioned analyses were added in response to pre-submission review and are post hoc. Analyses used Python 3.9.6 (package versions in Supplementary S1.14). The statistical significance threshold was *P* < .05.

### 2.6 Ethics

All data were de-identified and came from public research databases released under their own institutional ethics approvals, so no additional review board approval, informed consent or trial registration was required.

## 3. Results

### 3.1 Cohort

Of 6,009 intensive care stays with a cardiac arrest, anoxic or hypoxic-ischaemic diagnosis, 4,330 met the first-stay, disposition, named-arrest and timing criteria, and 3,641 of these had an APACHE IVa predicted hospital mortality and formed the primary cohort, drawn from 161 hospitals (Figure 1; Table S1).

**Figure 1.**
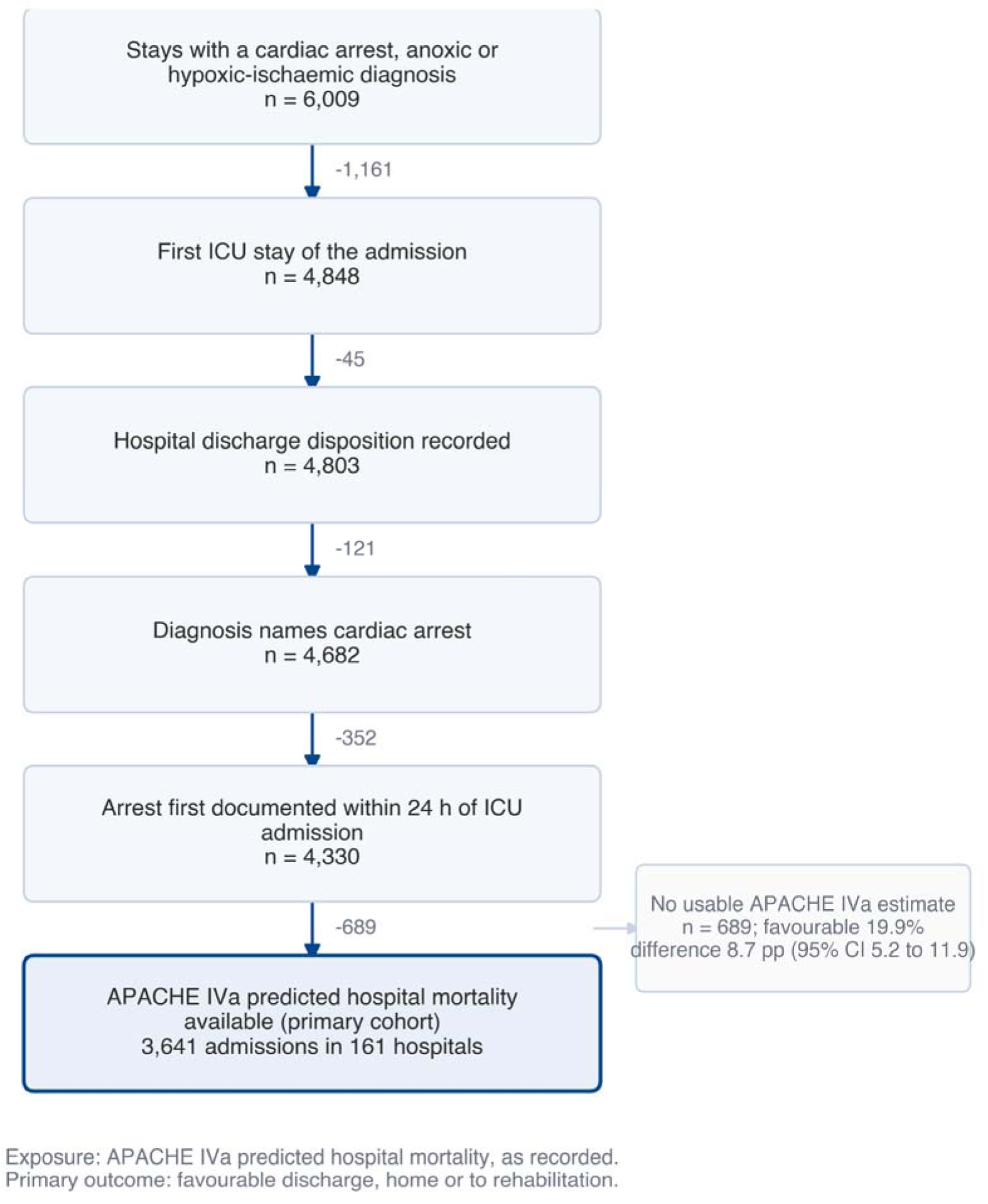
Cohort selection. Selection of the primary cohort from the eICU Collaborative Research Database. Counts to the right of each arrow are the admissions excluded at that step. Admissions without a usable APACHE IVa predicted hospital mortality are shown separately, because that exclusion shifted the outcome distribution: their favourable-discharge proportion was lower than that of the admissions retained, so the analytic cohort overstates favourable discharge relative to all candidate post-arrest admissions.

Median age was 65 years (IQR, 54 to 75), 2,107 patients (57.9%) were male, and 2,811 (77.2%) were mechanically ventilated. Median Glasgow Coma Scale motor response was 1; 1,791 patients (49.2%) had no motor response, and motor response was missing for 173 (4.8%). Median APACHE IVa predicted hospital mortality was 58.3% (IQR, 29.9 to 78.7). In hospital, 1,837 patients (50.5%) died and 1,039 (28.5%) were discharged home or to rehabilitation (Table 1).

**Table 1.** Characteristics of the primary cohort, split at the pre-specified high-risk line. Values are median (IQR) or No. (%). No hypothesis tests are reported, because APACHE IVa defines the strata. GCS, Glasgow Coma Scale; IQR, interquartile range.

| Characteristic | Overall (n=3,641) | Predicted mortality <50% (n=1,502) | Predicted mortality ≥50% (n=2,139) |
| --- | --- | --- | --- |
| Age, median (IQR), years | 65 (54 to 75) | 63 (52 to 73) | 67 (56 to 77) |
| Male sex, No. (%) | 2107 (57.9) | 893 (59.5) | 1214 (56.8) |
| GCS motor response, median (IQR) | 1 (1 to 6) | 6 (4 to 6) | 1 (1 to 1) |
| GCS total, median (IQR) | 3 (3 to 11) | 13 (7 to 15) | 3 (3 to 3) |
| Mechanically ventilated, No. (%) | 2811 (77.2) | 889 (59.2) | 1922 (89.9) |
| Intubated, No. (%) | 2103 (57.8) | 559 (37.2) | 1544 (72.2) |
| Arterial pH, median (IQR) | 7.32 (7.22 to 7.40) | 7.35 (7.29 to 7.41) | 7.31 (7.20 to 7.39) |
| Predicted hospital mortality, median (IQR), % | 58.3 (29.9 to 78.7) | 22.6 (9.1 to 37.8) | 75.7 (63.6 to 85.2) |
| Died in hospital, No. (%) | 1837 (50.5) | 395 (26.3) | 1442 (67.4) |
| Discharged home or to rehabilitation, No. (%) | 1039 (28.5) | 738 (49.1) | 301 (14.1) |

#### Requiring an APACHE IVa estimate selected the cohort

The 689 (15.9%) otherwise eligible admissions without a usable prediction had a lower favourable-discharge proportion than those retained (19.9% vs 28.5%; risk difference, 8.7 percentage points; 95% CI, 5.2 to 11.9) and a higher in-hospital mortality (58.2% vs 50.5%), so the analytic cohort overstates favourable discharge.

### 3.2 Favourable discharge declines steeply but remains observed at the highest predicted mortality

**Favourable discharge showed a strong downward trend across deciles of APACHE IVa-predicted hospital mortality**, from 78.9% in the lowest decile to 4.1% in the highest (Cochran-Armitage z, -27.2; P < 0.001; adjusted P < 0.001) (Figure 2a). The gradient held for discharge home only (z, -27.2) and for survival to discharge (z, -28.5).

**Figure 2.**
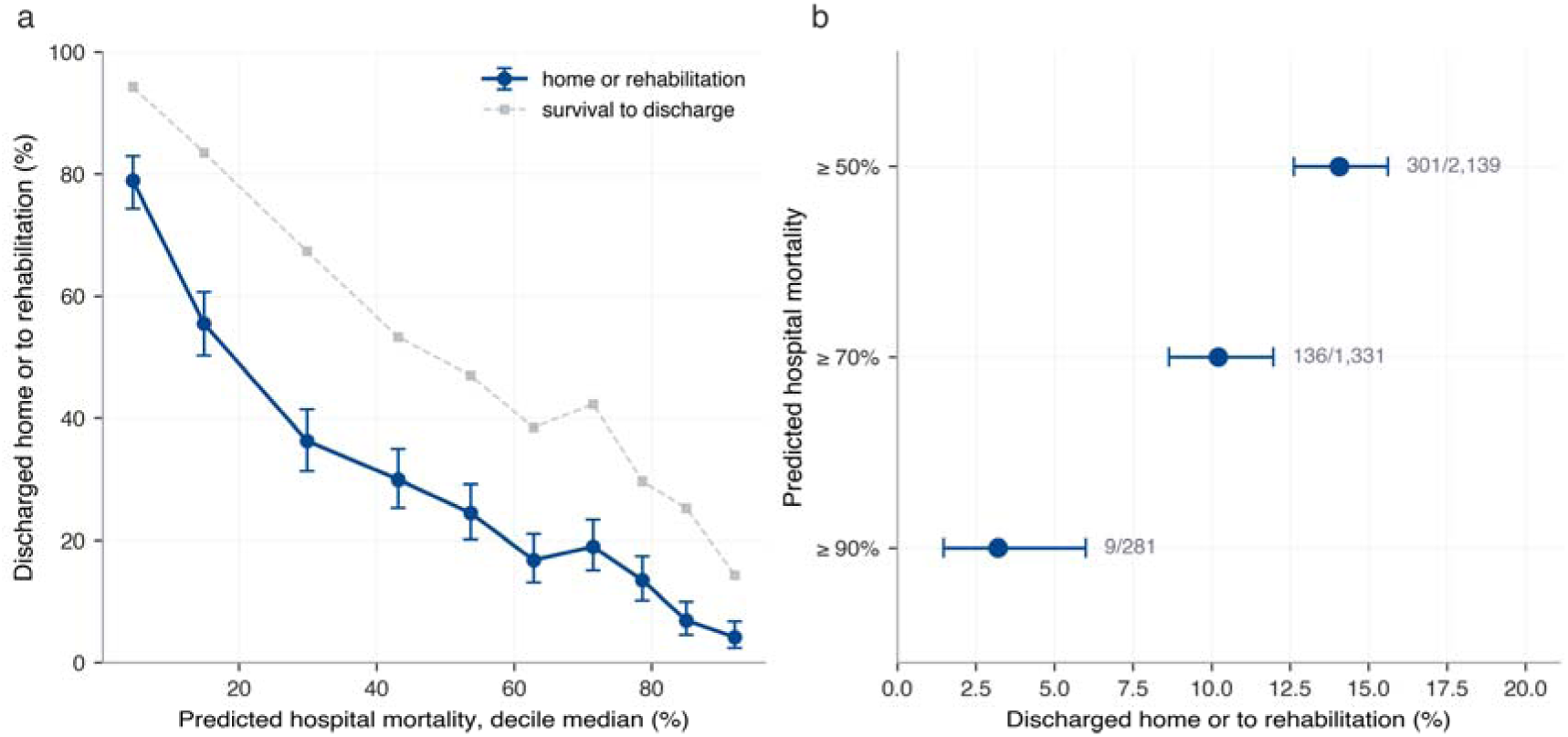
Favourable discharge across the range of APACHE IVa-predicted hospital mortality. (a) Proportion discharged home or to rehabilitation by decile of APACHE IVa predicted hospital mortality, plotted against the decile median, with exact 95% confidence intervals; survival to hospital discharge is shown for comparison. (b) Proportion discharged home or to rehabilitation at three thresholds of predicted mortality, with counts beside each interval. n = 3,641 admissions in 161 hospitals.

**Among the 2,139 patients whose APACHE IVa-predicted hospital mortality was at or above 50%, 301 (14.1%; exact 95% CI, 12.6 to 15.6; hospital-clustered 95% CI, 12.6 to 15.6) were discharged home or to rehabilitation** (Figure 2b), a proportion exceeding 5% (exact binomial P < 0.001; adjusted P < 0.001). Discharge home gave 248 (11.6%; 95% CI, 10.3 to 13.0), and survival to discharge 697 (32.6%; 95% CI, 30.6 to 34.6).

Favourable discharge persisted at more extreme values: 136 of 1,331 patients (10.2%; 95% CI, 8.6 to 12.0) with predicted mortality at or above 70%, and 9 of 281 (3.2%; 95% CI, 1.5 to 6.0) at or above 90% (Table S14).

### 3.3 No pre-specified subgroup met the low-recovery benchmark

**None of the 14 pre-specified bedside subgroups met the low-recovery benchmark** (Figure 3; Table S5). The lowest observed proportion was in patients with predicted mortality at or above 90% and no motor response, 8 of 264 (3.0%; 95% CI, 1.3 to 5.9): the point estimate fell below 5% but the exact upper bound did not, and the classification was unchanged on the hospital-clustered bound. At a more permissive 10% benchmark, 2 of 14 subgroups qualified. Within the high-risk stratum, favourable discharge ranged from 8.0% (69 of 860; 95% CI, 6.3 to 10.0) in patients 65 years or older with a motor response of 2 or less to 22.5% (64 of 285; 95% CI, 17.7 to 27.8) in patients younger than 50 years.

**Figure 3.**
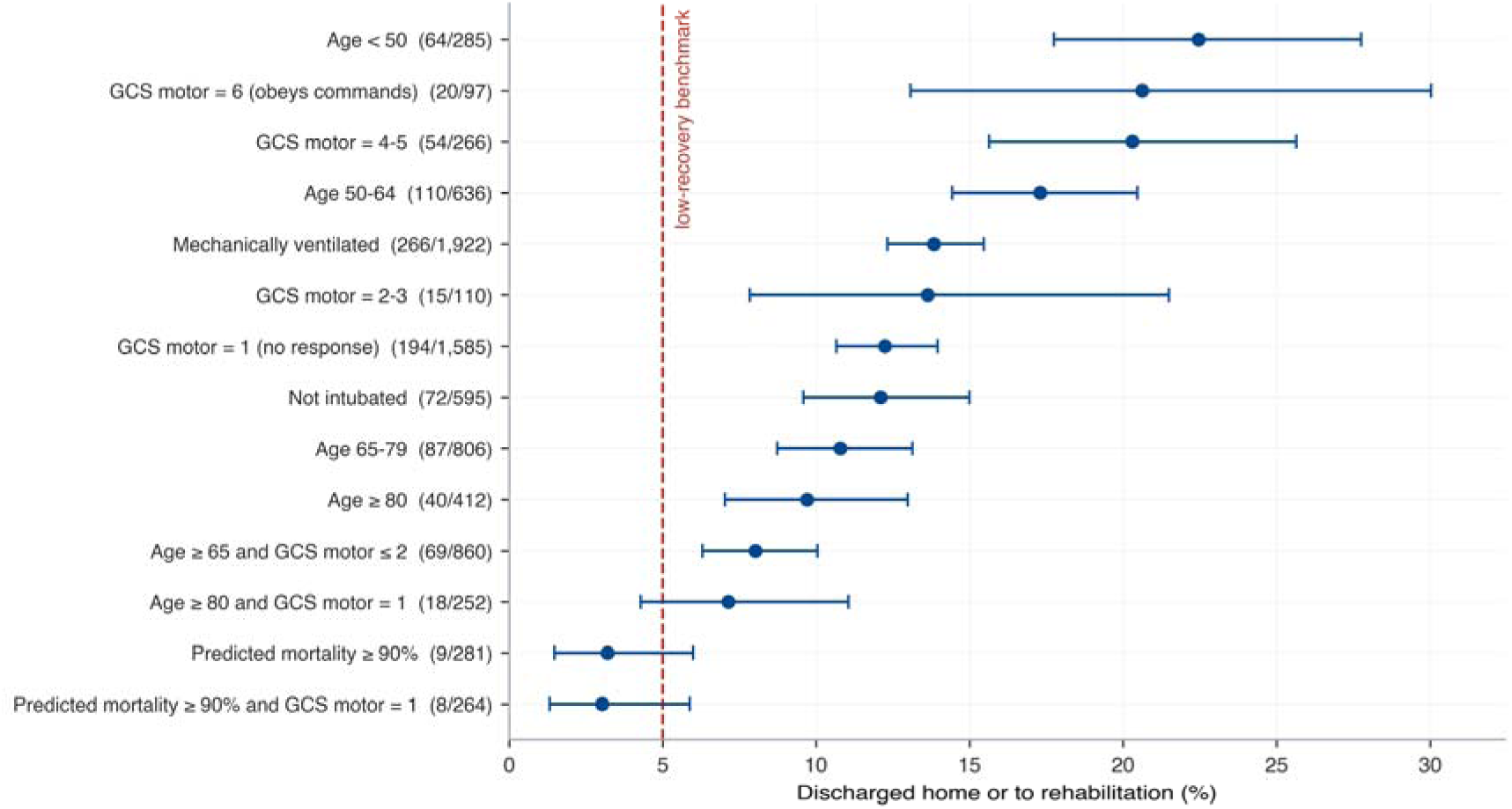
No pre-specified subgroup met the low-recovery benchmark. Proportion discharged home or to rehabilitation in each of 14 pre-specified bedside subgroups within the high-risk stratum (APACHE IVa-predicted hospital mortality at or above 50%), with exact 95% confidence intervals and counts. The dashed line marks the pre-specified low-recovery benchmark of 5%, an analytic benchmark rather than a clinical threshold; a subgroup would meet it only if its entire interval fell to the left. None did. Exact intervals are displayed; classification was also evaluated on the hospital-clustered upper bound and was unchanged. Subgroups overlap and are not mutually exclusive.

#### Age and motor response were both associated with favourable discharge after adjustment

In 2,058 high-risk patients across 152 hospitals, each additional 10 years of age was associated with lower odds (odds ratio, 0.78; 95% CI, 0.73 to 0.84) and each additional point of motor response with higher odds (odds ratio, 1.22; 95% CI, 1.10 to 1.35); male sex was weakly associated (odds ratio, 1.33; 95% CI, 1.02 to 1.73) and mechanical ventilation was not (odds ratio, 0.89; 95% CI, 0.57 to 1.39) (Table S8). The age trend across the four pre-specified bands was significant (Cochran-Armitage z, -5.70; P < 0.001; adjusted P < 0.001).

#### Favourable discharge remained observable in patients not obeying commands

Of the 1,961 high-risk patients not obeying commands during the APACHE observation period, 263 (13.4%; exact 95% CI, 11.9 to 15.0; clustered 95% CI, 12.0 to 14.8) had a favourable discharge.

### 3.4 Discrimination did not differ detectably between the two endpoints

The area under the curve was 0.789 (95% CI, 0.771 to 0.805) for unfavourable functional outcome and 0.774 (95% CI, 0.751 to 0.798) for in-hospital death. The difference, in-hospital death minus unfavourable functional outcome, was -0.015 (95% CI, -0.032 to 0.001; P = 0.078; adjusted P = 0.078) (Table S9). No equivalence margin was pre-specified, so the analysis supports only the absence of a detectable difference.

### 3.5 Independent cohort

In MIMIC-IV, 2,653 admissions with a documented cardiac-arrest code had a median age of 65 years, 56.8% died in hospital and 631 (23.8%) were discharged home, to home health care or to rehabilitation. MIMIC-IV records no APACHE score, so this examines the bedside gradients rather than replicating the APACHE result. Favourable discharge fell from 29.4% in patients younger than 50 years to 13.1% in those 80 years or older (Figure 4a), and from 36.5% in patients obeying commands to 9.6% in those with no motor response (Figure 4b); among patients 65 years or older with a motor response of 2 or less it was 5.7% (Table S10).

**Figure 4.**
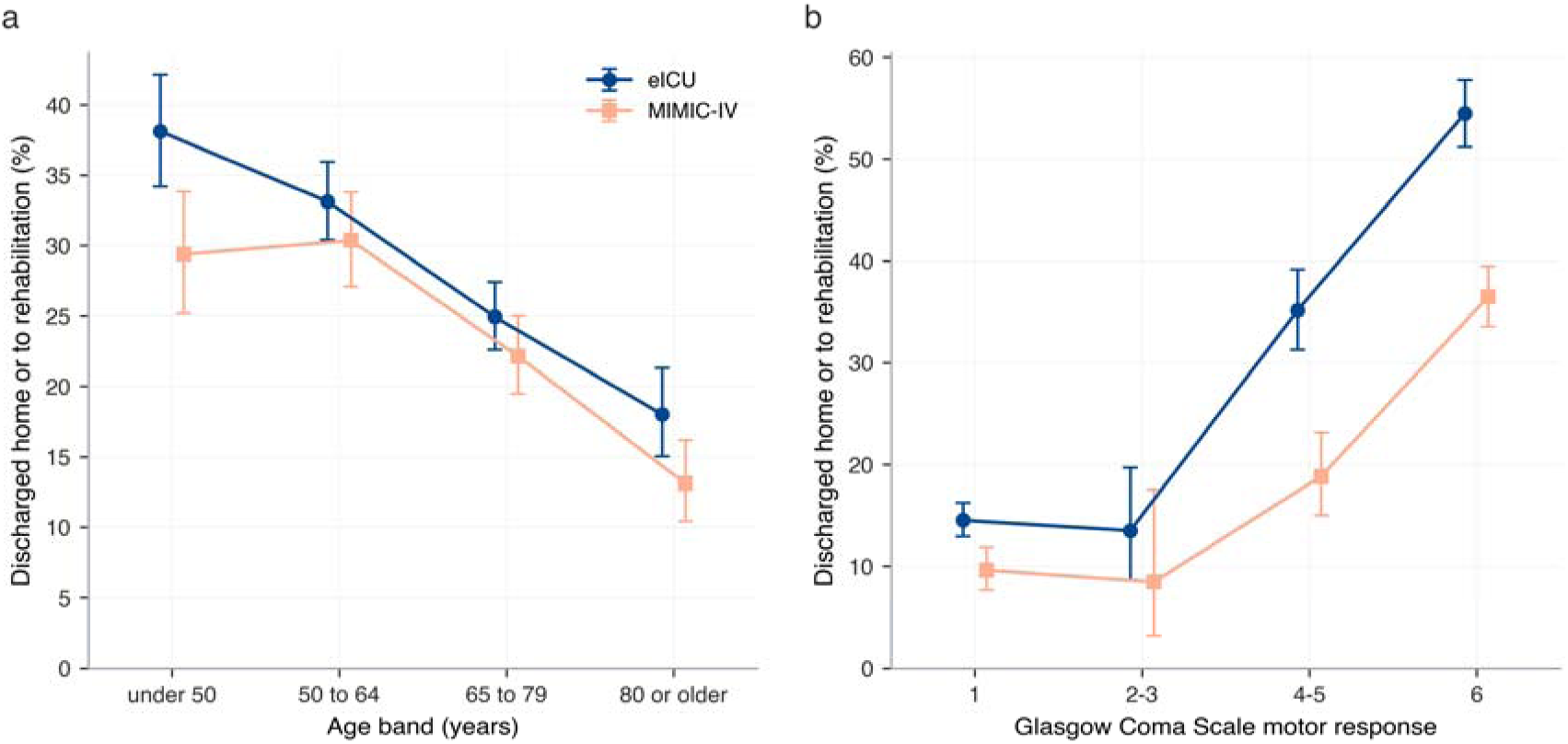
Independent replication of the bedside gradients. Proportion discharged home or to rehabilitation by (a) age band and (b) Glasgow Coma Scale motor response, in the full eICU analytic cohort and in the full MIMIC-IV post-arrest cohort, with exact 95% confidence interval. Band definitions are identical in the two cohorts, and both are shown whole rather than restricted to a severity stratum, so the two series describe the same populations. MIMIC-IV records no APACHE score, so this replicates the bedside gradients rather than the APACHE result; a motor band with fewer than 20 patients would be omitted; none was.

### 3.6 Secondary and Sensitivity Analyses

Under both alternative outcome definitions no subgroup met the benchmark (Tables S2 to S7). Favourable discharge in the high-risk stratum ranged from 12.1% to 15.3% across sensitivity analyses: 15.3% after excluding 174 inter-hospital transfers, 12.1% in the 595 patients not intubated, 14.3% and 13.3% with diagnosis-timing windows of 360 and 60 minutes, and 14.7% with eligibility anchored on the APACHE admission diagnosis; the three patient-level primary tests remained significant with hospital clustering (Supplementary S2.1). Of the 697 high-risk hospital survivors, 301 (43.2%; 95% CI, 39.5 to 47.0) had a favourable discharge (Supplementary S2.2). In I-CARE, an unlinked cohort, 225 of 607 comatose post-arrest patients (37.1%) reached Cerebral Performance Category 1 or 2 at 3 to 6 months (Supplementary S2.3; Figure S1).

## 4. Discussion

Across 3,641 post-cardiac-arrest intensive care admissions in 161 hospitals, favourable discharge fell steeply with APACHE IVa-predicted hospital mortality, yet about one in seven patients whose predicted mortality was at or above 50% were discharged home or to rehabilitation. None of 14 pre-specified bedside subgroups allowed that proportion to be bounded below 5%. The conclusion is benchmark-dependent: at a more permissive 10% benchmark, two subgroups qualified, both defined by a predicted mortality at or above 90%. Discrimination was broadly similar for the two endpoints, so this is not a failure of ranking. General illness severity and evidence that a favourable outcome is absent are different quantities.

The central interpretive constraint is that most in-hospital deaths after cardiac arrest follow withdrawal of life-sustaining therapy.^4,5^ Withdrawal was not observed in either database, so self-fulfilling-prophecy bias cannot be quantified here; the proportions we report may understate those that would have been observed under continued treatment. Cohorts assembled to be uncensored by withdrawal report higher survival with good function than registry averages,^4^ consistent with that direction, and current guidance cites this bias as a reason to avoid reliance on any single early predictor.^2^

Our results sit alongside purpose-built post-arrest scores rather than against them. The CAST score, developed for post-cardiac-arrest syndrome, reached a specificity of 1.00 in external validation at its most stringent cutoff and has since stratified severity in a multicentre registry of 1,111 patients.^8,9^ It draws on arrest characteristics, blood chemistry and brain imaging, most of which require deliberate collection and are not values a general severity score assembles. Such instruments may stratify post-arrest severity more usefully than a general ICU mortality estimate, and should still be read within a multimodal strategy.^2^

For the clinician, an APACHE IVa-predicted hospital mortality at or above 50% after cardiac arrest is compatible with a favourable-discharge proportion near one in seven, including among patients not obeying commands during the APACHE observation period, a subgroup that approximates but is not equivalent to the guideline population assessed at 72 hours or later. A general ICU mortality estimate is therefore not on its own evidence that functional recovery will not occur. These findings support the guideline position of multimodal prognostication at 72 hours or later.^2,3^

### 4.1 Limitations

First, discharge destination is a proxy for functional outcome that also reflects insurance status, bed availability and local practice; we carried three outcome definitions, excluded transfers in a sensitivity analysis and placed the proxy beside Cerebral Performance Category in a separate cohort, but no patient contributed to both. Second, requiring an APACHE IVa estimate excluded 689 admissions whose favourable-discharge proportion was 8.7 percentage points lower, so the analytic cohort overstates favourable discharge. Third, APACHE IVa physiology is derived from the first APACHE day, and we did not establish when or whether the estimate reached treating clinicians. Fourth, Glasgow Coma Scale values come from the APACHE physiology record rather than an earliest unsedated examination, so a motor score of 1 may reflect sedation, neuromuscular blockade or later deterioration; we repeated the analysis in patients not intubated. Fifth, withdrawal of life-sustaining therapy was not recorded, so its effect is argued in direction rather than quantified. Sixth, the public eICU release is a stratified sample, so no absolute rate is identifiable, and no post-discharge follow-up was available. Seventh, the eICU cohort dates from 2014 and 2015, and post-arrest management, withdrawal practice and rehabilitation pathways have changed since. Eighth, out-of-hospital and in-hospital arrest could not be reliably distinguished; admission source is only a proxy. Ninth, arrest rhythm, time to return of spontaneous circulation and targeted temperature management were not available in a form suitable for adjustment, which also prevented computing a purpose-built post-arrest score in the same patients.

### 4.2 Conclusion

Among post-cardiac-arrest intensive care admissions, favourable discharge declined markedly as APACHE IVa-predicted hospital mortality rose but remained observable at the highest risk levels, and no pre-specified bedside subgroup allowed it to be bounded below 5%, although two qualified at 10%. A general ICU mortality estimate should not be interpreted as standalone evidence that a favourable functional outcome is absent.

## Data Availability

eICU-CRD v2.0, MIMIC-IV v3.1 and I-CARE v2.1 are available on PhysioNet to credentialed users who complete the required training.

https://physionet.org/content/eicu-crd/2.0/

https://physionet.org/content/mimiciv/3.1/

https://physionet.org/content/i-care/2.1/

https://github.com/Alon-Gorenshtein/favourable_discharge_apache

## Declarations

## Ethics approval

All analyses used previously collected, fully de-identified data from public research databases distributed under their respective data-use agreements. Each source database was collected and released under its own institutional ethics approval. No patient contact and no new data collection occurred, so no additional institutional review board review or informed consent was required for this secondary analysis.

## Declaration of generative AI in the writing process

During the preparation of this work the authors used Claude (Anthropic) to assist with language editing, structural refinement, critical review of the manuscript, and checks of clarity and internal consistency, and some of the secondary analyses reported here were added in response to that review. All suggestions were independently evaluated by the authors, who performed and verified every analysis, reviewed and edited the final manuscript, and take full responsibility for its content.

## Data availability

eICU-CRD v2.0, MIMIC-IV v3.1 and I-CARE v2.1 are available on PhysioNet to credentialed users who complete the required training.^15^

## Code availability

Analysis code reproducing every reported number is available at https://github.com/Alon-Gorenshtein/favourable_discharge_apache.

## CRediT author statement

Author contributions are described using the CRediT (Contributor Roles Taxonomy) framework. **Alon Gorenshtein (A.G.):** conceptualization, methodology, software, formal analysis, data curation, validation, visualization, and writing of the original draft. **Amir Srour (A.S.):** review and editing of the manuscript. **Yosef Adiniaev (Y.A.):** software, data curation, formal analysis, validation, and review and editing of the manuscript. **Yiftach Barash (Y.B.):** methodology, validation, and review and editing of the manuscript. **Eyal Klang (E.K.):** conceptualization, methodology, supervision, resources, and review and editing of the manuscript. **Oved Daniel (O.D.):** conceptualization, clinical interpretation, supervision, and review and editing of the manuscript. All authors critically reviewed the manuscript, approved the final version submitted for publication, and agree to be accountable for all aspects of the work. The corresponding author (A.G.) had full access to all data in the study and takes responsibility for the integrity of the data and the accuracy of the analysis.

