## Supplementary Information for "Favourable discharge after cardiac arrest across APACHE IVa-predicted hospital mortality: a multicentre observational study"

This Supplementary Information accompanies the main manuscript. It contains the extended methods, the secondary and sensitivity results summarised in the main text, the cohort flow, the full per-decile and per-subgroup result tables under all three outcome definitions, the adjusted regression output, and the pre-specified test family with its multiplicity correction.

**Contents**

- S1 Supplementary Methods
- S2 Supplementary Results
- S3 Supplementary Tables (Table S1 to Table S14)
- S4 Supplementary Figure (Figure S1)
- S5 Code and Data Availability

### S1 Supplementary Methods

#### S1.1 Cohort construction

eICU-CRD v2.0 stays were retained when a recorded diagnosis string contained cardiac arrest, anoxic, or hypoxic-ischemic, matched case-insensitively against the full diagnosis path. The primary cohort also required that the diagnosis string name cardiac arrest, and that it was first documented no more than 1,440 minutes after intensive care admission. The stay had to be the patient's first, to carry a recorded hospital discharge disposition, and to carry an APACHE IVa predicted hospital mortality between 0 and 1.

The eICU diagnosis table is an active list, so a diagnosis may be entered at any point during the stay. Anchoring on the diagnosis offset ensures the arrest was documented before or early within the APACHE observation period rather than later in the stay. In the primary cohort the median offset was 86 minutes. An alternative anchor is the APACHE admission diagnosis, which eICU records as a field on the patient table rather than as a separate table; eligibility was re-derived from that field as a sensitivity analysis and is reported in S2.1.

Because the earlier version of this work used the broader definition, that cohort is retained as a sensitivity analysis and reported in S2.1.

MIMIC-IV v3.1 admissions were retained when a diagnosis carried an explicit cardiac-arrest code (ICD-9 4275, ICD-10 I46), matching the eICU requirement for a named arrest. Admissions carrying only an anoxic brain injury code (ICD-9 3481, ICD-10 G931) were analysed separately as a broader sensitivity cohort; admitting anoxic brain injury without a coded arrest, as an earlier version of this work did, gave 3,308 admissions with a favourable-discharge proportion of 23.9%. I-CARE v2.1 supplied 607 comatose post-arrest patients undergoing continuous electroencephalography, with Cerebral Performance Category assessed at 3 to 6 months after return of spontaneous circulation.

#### S1.2 Selection created by requiring an APACHE IVa estimate

APACHE predictions are not generated for every intensive care admission; stays shorter than four hours and several other categories are designated non-predictive. Of 4,330 otherwise eligible admissions, 689 had no usable APACHE IVa prediction. Those patients had a lower favourable-discharge proportion (19.9% versus 28.5%; risk difference 8.7 percentage points, 95% CI 5.2 to 11.9) and higher in-hospital mortality (58.2% versus 50.5%). The analytic cohort therefore overstates favourable discharge relative to all candidate post-arrest admissions, and the direction of that bias is stated in the main text.

#### S1.3 The APACHE IVa record

The apachePatientResult table carries both an APACHE IV and an APACHE IVa row for each stay. The extraction filters on the APACHE version field and retains only the IVa row before any deduplication, so no APACHE IV value enters the analysis. Predicted mortality values outside the interval 0 to 1 were discarded.

APACHE IVa physiology is derived from the first APACHE day rather than an admission-time snapshot. This study describes a quantity recorded in the source database; it does not establish when, or whether, that quantity was available to treating clinicians.

#### S1.4 The sampling constraint on the eICU release

The public eICU-CRD release is a stratified random sample of the underlying eICU Research Institute repository, stratified by hospital, and the per-hospital sampling fractions are not published. Any quantity expressed per hospital per unit time is therefore not identifiable. Proportions and associations within the sample are unaffected. Every quantity reported in this work is a proportion, an odds ratio, an area under a curve, or a trend statistic. No accrual rate, no per-hospital-year count, and no absolute time appears anywhere in the manuscript or in this supplement.

#### S1.5 Outcome definitions

The primary outcome was favourable discharge disposition, defined as discharge home or to rehabilitation. In eICU this was the discharge-location values Home and Rehabilitation. In MIMIC-IV it was HOME, HOME HEALTH CARE, and REHAB. All other dispositions, including skilled nursing facility, long-term acute care, transfer to another hospital, hospice, and death in hospital, were unfavourable. Because the final outcome of patients transferred to another acute hospital is unknown, a sensitivity analysis excluded them rather than classing them unfavourable.

Two further outcome definitions were carried through every analysis: discharge home only, and survival to hospital discharge. The three are nested in that order.

#### S1.6 The low-recovery benchmark

The rule was fixed before subgroup results were generated. A subgroup met the benchmark only when the upper limit of the 95% confidence interval on its favourable-discharge proportion fell strictly below 0.05. The point estimate carried no weight.

The 5% value is an analytic benchmark, not a clinical threshold for withdrawal of life-sustaining therapy. No clinical consensus establishes that a 4.9% probability of favourable outcome is meaningfully different from 5.1% for a treatment decision. The classification is reported at 5% and at a more permissive 10%, on both the exact and the hospital-clustered interval. Because the rule tests an upper confidence bound, failure at 5% entails failure at any stricter benchmark, so 2.5% and 1% are not independent sensitivity analyses. These are per-subgroup confidence bounds and do not provide simultaneous family-wise coverage across the 14 pre-specified subgroups.

In this cohort the rule bound in exactly the way it was designed to. The lowest observed subgroup, patients with predicted mortality at or above 90% and no motor response, had 8 favourable discharges among 264, a point estimate of 3.0% that falls below the line. Its exact upper bound was 5.9%, so the group did not meet the benchmark. Reporting the point estimate alone would have reversed that conclusion.

A subgroup was analysed only when it contained at least 20 patients.

#### S1.7 Clustered uncertainty

Patients are clustered within hospitals and may share post-arrest treatment, withdrawal practice, rehabilitation access and discharge conventions. Exact binomial intervals treat patients as independent and therefore understate uncertainty. Every primary proportion also carries a percentile interval from a nonparametric bootstrap resampling hospitals with replacement over 5,000 replicates. Both intervals are reported. Clopper-Pearson intervals are conservative under the independent-binomial model. Because patients are clustered within hospitals that guarantee does not transfer to the actual sampling process, which is why the clustered interval is reported alongside rather than in place of it. No conclusion in this work differs between the two.

#### S1.8 Adjusted model

Within the high-risk stratum, favourable discharge was modelled on age in decades, sex, Glasgow Coma Scale motor response and mechanical ventilation, using generalised estimating equations with a binomial family, a logit link, and an exchangeable working correlation with hospital as the cluster. Patients missing any covariate were dropped, leaving 2,058 patients in 152 hospitals. These four coefficients are exploratory and their P values are not adjusted for multiplicity.

#### S1.9 Discrimination comparison

APACHE IVa predicted hospital mortality was evaluated as a ranking score for two endpoints in the same patients: in-hospital death, which the score was built to predict, and unfavourable functional outcome. The reported difference is the area for in-hospital death minus the area for unfavourable functional outcome. Confidence intervals came from a bootstrap over 2,000 replicates resampling hospitals rather than patients. No equivalence margin was pre-specified, so the analysis can support only the absence of a detectable difference, not equivalence.

#### S1.10 Multiplicity

Four primary tests were named before analysis: the trend across deciles, the comparison of high-risk favourable discharge against the 5% benchmark, the difference in discrimination between endpoints, and the trend across age bands within the high-risk stratum. These four were corrected together by the Benjamini-Hochberg procedure. No other family is corrected; the subgroup search is an interval-based classification rather than a hypothesis test, and the four regression coefficients are exploratory.

#### S1.11 Secondary sensitivity analyses

The following were carried out to support the primary result and were not part of the pre-specified multiplicity family. Diagnosis documentation was restricted to 360 and to 60 minutes after intensive care admission. Cohort eligibility was anchored on the APACHE admission diagnosis recorded in the patient table. Patients were stratified by admission source as a proxy for arrest setting. Inter-hospital transfers were excluded rather than counted unfavourable. Glasgow Coma Scale analyses were repeated in patients who were not intubated. The three patient-level primary tests were repeated with hospital-clustered generalised estimating equations. Favourable discharge was described conditional on survival to hospital discharge, which changes the estimand and is reported as secondary. The timing, admission-source, admission-diagnosis and survivor-conditioned analyses were added in response to pre-submission review and are post hoc.

#### S1.12 Variable encoding and threshold choice

The eICU patient table stores ages above 89 years as the string "> 89" rather than an exact value. These were encoded as 90 years. Because age enters the adjusted model in decades, this affects only the oldest band and cannot alter its ordering.

The high-risk threshold of 50% was chosen a priori as an intuitive boundary at which predicted mortality equals predicted survival. It was not intended as a clinical treatment threshold, and results are also reported at 70% and 90%.

#### S1.13 Data era

The primary eICU cohort is drawn from admissions in 2014 and 2015. Post-arrest management, targeted temperature management, withdrawal practice, rehabilitation access and discharge pathways have changed since, so absolute favourable-discharge proportions may differ in contemporary practice. The MIMIC-IV cohort spans a later and wider period but does not carry an APACHE score, so it does not resolve this directly.

#### S1.14 Pre-specification and software

The design document and implementation plan were committed to the authors' version-controlled working repository before the subgroup results were generated, at commits fc10472 and b526ffe respectively. That repository holds the manuscript sources alongside the code and is not public; the public code repository below is a code-only export, so these identifiers do not resolve within it. The plan documents are available from the corresponding author on request. Analyses used Python 3.9.6 with pandas 2.3.3, numpy 2.0.2, scipy 1.13.1, statsmodels 0.14.6, scikit-learn 1.6.1 and matplotlib 3.9.4. The random seed was fixed at 20260830 for every resampling procedure.

### S2 Supplementary Results

The main text summarises these analyses in Section 3.6; the full estimates are given here.

#### S2.1 Sensitivity analyses

Results were similar under both alternative outcome definitions, and the decile trend held under each (Tables S2 to S4). No subgroup met the benchmark under any of the three outcome definitions (Tables S5 to S7).

Excluding the 174 high-risk patients transferred to another acute hospital, whose final outcome is unknown, raised favourable discharge from 14.1% to 15.3% (301 of 1,965; 95% CI, 13.8 to 17.0). Restricting the high-risk stratum to the 595 patients not intubated, in whom the Glasgow Coma Scale is most interpretable, gave 72 favourable discharges (12.1%; 95% CI, 9.6 to 15.0). Motor response was missing for 173 of 3,641 patients (4.8%), who were excluded from motor-based subgroups and from the adjusted model, which used 2,058 of 2,139 high-risk patients. Tightening the diagnosis-timing window, which reduces the chance that eligibility reflected an arrest after intensive care admission, did not move the estimate: at 360 minutes, 277 of 1,943 (14.3%; 95% CI, 12.7 to 15.9); at 60 minutes, 133 of 1,003 (13.3%; 95% CI, 11.2 to 15.5), against 14.1% in the primary 1,440-minute cohort.

Refitting the three patient-level primary tests with hospital-clustered generalised estimating equations reached the same conclusions: the decile trend, the comparison of the high-risk stratum against the 5% benchmark and the age-band trend all remained significant (P < 0.001 for each). Anchoring eligibility on the APACHE admission diagnosis recorded in the patient table rather than on the diagnosis list gave a cohort of 2,961 admissions in which 285 of 1,939 (14.7%; 95% CI, 13.2 to 16.4) high-risk patients had a favourable discharge. Admission source, a proxy for where the arrest occurred, did not separate the result: 184 of 1,370 (13.4%) admitted from the emergency department and 69 of 445 (15.5%) from an inpatient location; this is a proxy, not a validated out-of-hospital versus in-hospital classification. Repeating the primary analysis on the broader cohort used in an earlier version of this work, which admitted anoxic and hypoxic-ischaemic diagnoses without a named arrest and applied no timing restriction, gave 308 of 2,208 (13.9%), within 0.2 percentage points of the primary estimate.

#### S2.2 Favourable discharge among hospital survivors

To separate functional disposition from the mortality component of the endpoint, we examined favourable discharge among hospital survivors. Of the 697 patients who survived despite a predicted hospital mortality at or above 50%, 301 (43.2%; 95% CI, 39.5 to 47.0) went home or to rehabilitation; of the 37 survivors whose predicted mortality was at or above 90%, 9 (24.3%; 95% CI, 11.8 to 41.2) did. Conditioning on survival changes the estimand and is reported as a secondary analysis.

#### S2.3 Outcome anchor in I-CARE

In I-CARE, where Cerebral Performance Category was assessed prospectively at 3 to 6 months after return of spontaneous circulation, 225 of 607 comatose post-arrest patients undergoing continuous electroencephalography (37.1%; 95% CI, 33.2 to 41.0) reached Category 1 or 2 (Figure S1; Tables S11 and S12), against a favourable proportion under the discharge-destination definition of 28.5% in eICU and 23.8% in MIMIC-IV. The cohorts differ in case mix, the outcomes are assessed at different times, and no patient was linked across definitions, so this comparison locates the proxy rather than validating it.

### S3 Supplementary Tables

**Table S1.** Cohort flow. Percentages are of the patients remaining at that step.

| Step | n | Favourable outcome (%) | Died in hospital (%) |
| --- | --- | --- | --- |
| Stays with a cardiac arrest, anoxic or hypoxic-ischaemic diagnosis | 6009 | 26.9 | 48.3 |
| First ICU stay of the admission | 4848 | 26.7 | 50.2 |
| Hospital discharge disposition recorded | 4803 | 27 | 50.7 |
| Diagnosis names cardiac arrest | 4682 | 27 | 51.2 |
| Arrest first documented within 24 h of ICU admission | 4330 | 27.2 | 51.7 |
| APACHE IVa predicted hospital mortality available (primary cohort) | 3641 | 28.5 | 50.5 |

**Table S2.** Favourable outcome by decile of predicted hospital mortality, under the definition discharge home or to rehabilitation.

| Decile | n | Predicted mortality, decile median (%) | No. favourable | Favourable outcome (%) | 95% CI lower | 95% CI upper | Died in hospital (%) |
| --- | --- | --- | --- | --- | --- | --- | --- |
| 1 | 365 | 4.5 | 288 | 78.9 | 74.4 | 83 | 5.8 |
| 2 | 364 | 14.9 | 202 | 55.5 | 50.2 | 60.7 | 16.5 |
| 3 | 364 | 29.9 | 132 | 36.3 | 31.3 | 41.4 | 32.7 |
| 4 | 364 | 43.1 | 109 | 29.9 | 25.3 | 34.9 | 46.7 |
| 5 | 364 | 53.7 | 89 | 24.5 | 20.1 | 29.2 | 53 |
| 6 | 364 | 62.8 | 61 | 16.8 | 13.1 | 21 | 61.5 |
| 7 | 364 | 71.5 | 69 | 19 | 15.1 | 23.4 | 57.7 |
| 8 | 364 | 78.7 | 49 | 13.5 | 10.1 | 17.4 | 70.3 |
| 9 | 364 | 85 | 25 | 6.9 | 4.5 | 10 | 74.7 |
| 10 | 364 | 92.1 | 15 | 4.1 | 2.3 | 6.7 | 85.7 |

**Table S3.** Favourable outcome by decile of predicted hospital mortality, under the definition discharge home only.

| Decile | n | Predicted mortality, decile median (%) | No. favourable | Favourable outcome (%) | 95% CI lower | 95% CI upper | Died in hospital (%) |
| --- | --- | --- | --- | --- | --- | --- | --- |
| 1 | 365 | 4.5 | 280 | 76.7 | 72 | 81 | 5.8 |
| 2 | 364 | 14.9 | 185 | 50.8 | 45.6 | 56.1 | 16.5 |
| 3 | 364 | 29.9 | 121 | 33.2 | 28.4 | 38.3 | 32.7 |
| 4 | 364 | 43.1 | 92 | 25.3 | 20.9 | 30.1 | 46.7 |
| 5 | 364 | 53.7 | 80 | 22 | 17.8 | 26.6 | 53 |
| 6 | 364 | 62.8 | 48 | 13.2 | 9.9 | 17.1 | 61.5 |
| 7 | 364 | 71.5 | 56 | 15.4 | 11.8 | 19.5 | 57.7 |
| 8 | 364 | 78.7 | 36 | 9.9 | 7 | 13.4 | 70.3 |
| 9 | 364 | 85 | 22 | 6 | 3.8 | 9 | 74.7 |
| 10 | 364 | 92.1 | 12 | 3.3 | 1.7 | 5.7 | 85.7 |

**Table S4.** Favourable outcome by decile of predicted hospital mortality, under the definition survival to hospital discharge.

| Decile | n | Predicted mortality, decile median (%) | No. favourable | Favourable outcome (%) | 95% CI lower | 95% CI upper | Died in hospital (%) |
| --- | --- | --- | --- | --- | --- | --- | --- |
| 1 | 365 | 4.5 | 344 | 94.2 | 91.3 | 96.4 | 5.8 |
| 2 | 364 | 14.9 | 304 | 83.5 | 79.3 | 87.2 | 16.5 |
| 3 | 364 | 29.9 | 245 | 67.3 | 62.2 | 72.1 | 32.7 |
| 4 | 364 | 43.1 | 194 | 53.3 | 48 | 58.5 | 46.7 |
| 5 | 364 | 53.7 | 171 | 47 | 41.8 | 52.2 | 53 |
| 6 | 364 | 62.8 | 140 | 38.5 | 33.4 | 43.7 | 61.5 |
| 7 | 364 | 71.5 | 154 | 42.3 | 37.2 | 47.6 | 57.7 |
| 8 | 364 | 78.7 | 108 | 29.7 | 25 | 34.7 | 70.3 |
| 9 | 364 | 85 | 92 | 25.3 | 20.9 | 30.1 | 74.7 |
| 10 | 364 | 92.1 | 52 | 14.3 | 10.9 | 18.3 | 85.7 |

**Table S5.** Every pre-specified bedside subgroup within the high-risk stratum (APACHE IVa-predicted hospital mortality at or above 50%) under the outcome definition home or rehabilitation (primary), ordered by observed proportion. Classification was evaluated on both the exact and the hospital-clustered upper 95% bound. Bounds are per-subgroup and do not provide simultaneous family-wise coverage across the 14 subgroups.

| Subgroup | n | No. favourable | Favourable (%) | Exact 95% CI | Clustered upper | Meets 5% | Meets 10% |
| --- | --- | --- | --- | --- | --- | --- | --- |
| predicted mortality 0.9 or above and motor 1 | 264 | 8 | 3 | 1.3 to 5.9 | 5.2 | No | Yes |
| predicted mortality 0.9 or above | 281 | 9 | 3.2 | 1.5 to 6.0 | 5.3 | No | Yes |
| age 80 or older and motor 1 | 252 | 18 | 7.1 | 4.3 to 11.1 | 10.3 | No | No |
| age 65 or older and motor 2 or less | 860 | 69 | 8 | 6.3 to 10.0 | 9.7 | No | No |
| age 80 or older | 412 | 40 | 9.7 | 7.0 to 13.0 | 13 | No | No |
| age 65 to 79 | 806 | 87 | 10.8 | 8.7 to 13.1 | 13 | No | No |
| not intubated | 595 | 72 | 12.1 | 9.6 to 15.0 | 15.1 | No | No |
| motor 1 (no response) | 1585 | 194 | 12.2 | 10.7 to 14.0 | 13.9 | No | No |
| motor 2 or 3 | 110 | 15 | 13.6 | 7.8 to 21.5 | 22 | No | No |
| mechanically ventilated | 1922 | 266 | 13.8 | 12.3 to 15.5 | 15.3 | No | No |
| age 50 to 64 | 636 | 110 | 17.3 | 14.4 to 20.5 | 20.5 | No | No |
| motor 4 or 5 | 266 | 54 | 20.3 | 15.6 to 25.6 | 25.5 | No | No |
| motor 6 (obeys commands) | 97 | 20 | 20.6 | 13.1 to 30.0 | 30.3 | No | No |
| age under 50 | 285 | 64 | 22.5 | 17.7 to 27.8 | 27.4 | No | No |

**Table S6.** Every pre-specified bedside subgroup within the high-risk stratum (APACHE IVa-predicted hospital mortality at or above 50%) under the outcome definition home only, ordered by observed proportion. Classification was evaluated on both the exact and the hospital-clustered upper 95% bound. Bounds are per-subgroup and do not provide simultaneous family-wise coverage across the 14 subgroups.

| Subgroup | n | No. favourable | Favourable (%) | Exact 95% CI | Clustered upper | Meets 5% | Meets 10% |
| --- | --- | --- | --- | --- | --- | --- | --- |
| predicted mortality 0.9 or above | 281 | 7 | 2.5 | 1.0 to 5.1 | 4.5 | No | Yes |
| predicted mortality 0.9 or above and motor 1 | 264 | 7 | 2.7 | 1.1 to 5.4 | 4.8 | No | Yes |
| age 65 or older and motor 2 or less | 860 | 57 | 6.6 | 5.1 to 8.5 | 8.4 | No | Yes |
| age 80 or older and motor 1 | 252 | 17 | 6.7 | 4.0 to 10.6 | 9.9 | No | No |
| age 65 to 79 | 806 | 68 | 8.4 | 6.6 to 10.6 | 10.6 | No | No |
| age 80 or older | 412 | 36 | 8.7 | 6.2 to 11.9 | 12.1 | No | No |
| motor 1 (no response) | 1585 | 162 | 10.2 | 8.8 to 11.8 | 12 | No | No |
| not intubated | 595 | 63 | 10.6 | 8.2 to 13.3 | 13.6 | No | No |
| mechanically ventilated | 1922 | 219 | 11.4 | 10.0 to 12.9 | 13 | No | No |
| motor 2 or 3 | 110 | 13 | 11.8 | 6.4 to 19.4 | 19.2 | No | No |
| age 50 to 64 | 636 | 91 | 14.3 | 11.7 to 17.3 | 17.4 | No | No |
| motor 4 or 5 | 266 | 41 | 15.4 | 11.3 to 20.3 | 19.7 | No | No |
| age under 50 | 285 | 53 | 18.6 | 14.3 to 23.6 | 22.8 | No | No |
| motor 6 (obeys commands) | 97 | 18 | 18.6 | 11.4 to 27.7 | 28.4 | No | No |

**Table S7.** Every pre-specified bedside subgroup within the high-risk stratum (APACHE IVa-predicted hospital mortality at or above 50%) under the outcome definition survival to hospital discharge, ordered by observed proportion. Classification was evaluated on both the exact and the hospital-clustered upper 95% bound. Bounds are per-subgroup and do not provide simultaneous family-wise coverage across the 14 subgroups.

| Subgroup | n | No. favourable | Favourable (%) | Exact 95% CI | Clustered upper | Meets 5% | Meets 10% |
| --- | --- | --- | --- | --- | --- | --- | --- |
| predicted mortality 0.9 or above and motor 1 | 264 | 33 | 12.5 | 8.8 to 17.1 | 17.1 | No | No |
| predicted mortality 0.9 or above | 281 | 37 | 13.2 | 9.4 to 17.7 | 17.6 | No | No |
| age 80 or older and motor 1 | 252 | 58 | 23 | 18.0 to 28.7 | 28.6 | No | No |
| age 65 or older and motor 2 or less | 860 | 227 | 26.4 | 23.5 to 29.5 | 30.4 | No | No |
| age 80 or older | 412 | 117 | 28.4 | 24.1 to 33.0 | 33.1 | No | No |
| motor 1 (no response) | 1585 | 456 | 28.8 | 26.6 to 31.1 | 32.5 | No | No |
| not intubated | 595 | 180 | 30.3 | 26.6 to 34.1 | 35.2 | No | No |
| age 50 to 64 | 636 | 202 | 31.8 | 28.2 to 35.5 | 36 | No | No |
| mechanically ventilated | 1922 | 614 | 31.9 | 29.9 to 34.1 | 35.1 | No | No |
| age 65 to 79 | 806 | 264 | 32.8 | 29.5 to 36.1 | 36.6 | No | No |
| motor 2 or 3 | 110 | 43 | 39.1 | 29.9 to 48.9 | 50 | No | No |
| age under 50 | 285 | 114 | 40 | 34.3 to 45.9 | 46.2 | No | No |
| motor 6 (obeys commands) | 97 | 43 | 44.3 | 34.2 to 54.8 | 53.4 | No | No |
| motor 4 or 5 | 266 | 127 | 47.7 | 41.6 to 53.9 | 53 | No | No |

**Table S8.** Adjusted association between bedside variables and favourable outcome within the high-risk stratum. Generalised estimating equations, binomial family, exchangeable working correlation clustered on hospital; 2058 patients in 152 hospitals with complete covariates.

| Variable | Odds ratio | 95% CI lower | 95% CI upper | P |
| --- | --- | --- | --- | --- |
| age (per 10 years) | 0.78 | 0.73 | 0.84 | <0.001 |
| male sex | 1.33 | 1.02 | 1.73 | 0.034 |
| GCS motor (per point) | 1.22 | 1.1 | 1.35 | <0.001 |
| mechanical ventilation | 0.89 | 0.57 | 1.39 | 0.608 |

**Table S9.** Discrimination of APACHE IVa predicted hospital mortality for the endpoint it was designed to predict and for unfavourable functional outcome, in the same patients. Confidence intervals from a bootstrap resampling hospitals rather than patients.

| endpoint | auroc | ci_lo | ci_hi |
| --- | --- | --- | --- |
| in-hospital death (designed) | 0.774 | 0.751 | 0.798 |
| unfavourable functional outcome | 0.789 | 0.771 | 0.805 |

**Table S10.** Replication in MIMIC-IV. This database records no APACHE score, so strata are formed from the two bedside variables that carried the association in the primary cohort.

| stratum | n | n_fav | fav | lo | hi |
| --- | --- | --- | --- | --- | --- |
| age under 50 | 446 | 131 | 29.4 | 25.2 | 33.8 |
| age 50 to 64 | 751 | 228 | 30.4 | 27.1 | 33.8 |
| age 65 to 79 | 899 | 199 | 22.1 | 19.5 | 25 |
| age 80 or older | 557 | 73 | 13.1 | 10.4 | 16.2 |
| motor 1 | 790 | 76 | 9.6 | 7.7 | 11.9 |
| motor 2 or 3 | 71 | 6 | 8.5 | 3.2 | 17.5 |
| motor 4 or 5 | 372 | 70 | 18.8 | 15 | 23.2 |
| motor 6 | 1058 | 386 | 36.5 | 33.6 | 39.5 |
| age 65 or older and motor 2 or less | 384 | 22 | 5.7 | 3.6 | 8.5 |

**Table S11.** Favourable-outcome proportion in each cohort under its own outcome definition. No patient contributed to more than one definition, so this locates the discharge-destination proxy beside the reference outcome rather than validating it within a patient.

| cohort | outcome_definition | n | n_favourable | favourable_rate | lo | hi |
| --- | --- | --- | --- | --- | --- | --- |
| eICU | discharge home or rehabilitation | 3641 | 1039 | 28.5 | 27.1 | 30 |
| MIMIC-IV | discharge home, home health or rehabilitation | 2653 | 631 | 23.8 | 22.2 | 25.5 |
| I-CARE | Cerebral Performance Category 1 or 2 | 607 | 225 | 37.1 | 33.2 | 41 |

**Table S12.** Distribution of Cerebral Performance Category at follow-up in the I-CARE cohort. Categories 1 and 2 form the favourable group.

| Cerebral Performance Category | n | Proportion (%) |
| --- | --- | --- |
| 1 | 181 | 29.8 |
| 2 | 44 | 7.2 |
| 3 | 20 | 3.3 |
| 4 | 9 | 1.5 |
| 5 | 353 | 58.2 |

**Table S13.** The four pre-specified primary tests, with Benjamini-Hochberg adjustment across the family.

| ID | Test | Method | Statistic | P | Adjusted P |
| --- | --- | --- | --- | --- | --- |
| T1 | Trend in favourable outcome across predicted-mortality deciles | Cochran-Armitage | -27.186 | <0.001 | <0.001 |
| T2 | Favourable outcome in the high-risk stratum exceeds 5% | exact binomial, one-sided | 0.141 | <0.001 | <0.001 |
| T3 | Discrimination differs between the designed and functional endpoints | paired cluster bootstrap | -0.015 | 0.078 | 0.078 |
| T4 | Trend in favourable outcome across age bands within the high-risk stratum | Cochran-Armitage | -5.702 | <0.001 | <0.001 |

**Table S14.** Favourable outcome at three thresholds of APACHE IVa-predicted hospital mortality, under each of the three pre-specified outcome definitions. Exact intervals are Clopper-Pearson; hospital-clustered intervals are shown for the primary outcome only.

| Outcome definition | Predicted hospital mortality | n | No. favourable | Favourable, % (exact 95% CI) | Clustered 95% CI |
| --- | --- | --- | --- | --- | --- |
| Home or rehabilitation | ≥50% | 2,139 | 301 | 14.1 (12.6 to 15.6) | 12.6 to 15.6 |
| Home or rehabilitation | ≥70% | 1,331 | 136 | 10.2 (8.6 to 12.0) | 8.7 to 11.8 |
| Home or rehabilitation | ≥90% | 281 | 9 | 3.2 (1.5 to 6.0) | 1.5 to 5.3 |
| Home only | ≥50% | 2,139 | 248 | 11.6 (10.3 to 13.0) |  |
| Home only | ≥70% | 1,331 | 108 | 8.1 (6.7 to 9.7) |  |
| Home only | ≥90% | 281 | 7 | 2.5 (1.0 to 5.1) |  |
| Survival to hospital discharge | ≥50% | 2,139 | 697 | 32.6 (30.6 to 34.6) |  |
| Survival to hospital discharge | ≥70% | 1,331 | 356 | 26.7 (24.4 to 29.2) |  |
| Survival to hospital discharge | ≥90% | 281 | 37 | 13.2 (9.4 to 17.7) |  |

### S4 Supplementary Figure

**Figure S1. Outcome anchor.** Favourable-outcome proportion in each cohort under its own outcome definition, with exact 95% confidence intervals. eICU and MIMIC-IV use discharge destination; I-CARE uses Cerebral Performance Category 1 or 2, assessed prospectively at 3 to 6 months after return of spontaneous circulation. The cohorts differ in case mix, the outcomes are assessed at different times, and no patient contributed to more than one definition, so this locates the discharge-destination proxy beside a reference outcome rather than validating it within a patient. It is reported in the supplement for that reason.

### S5 Code and Data Availability

eICU-CRD v2.0, MIMIC-IV v3.1 and I-CARE v2.1 are distributed by PhysioNet to credentialed users who have completed the required human-subjects training. This study added no new data collection.

Analysis code reproducing every number in the manuscript and in this supplement from the source databases is available at <https://github.com/Alon-Gorenshtein/favourable_discharge_apache>. Each reported quantity is written to a machine-readable results digest by the analysis scripts and read from that digest by the manuscript, so no number is transcribed by hand.
