## Supplementary figures and images for "Favourable discharge after cardiac arrest across APACHE IVa-predicted hospital mortality: a multicentre observational study"

### Supplemental Figure S1

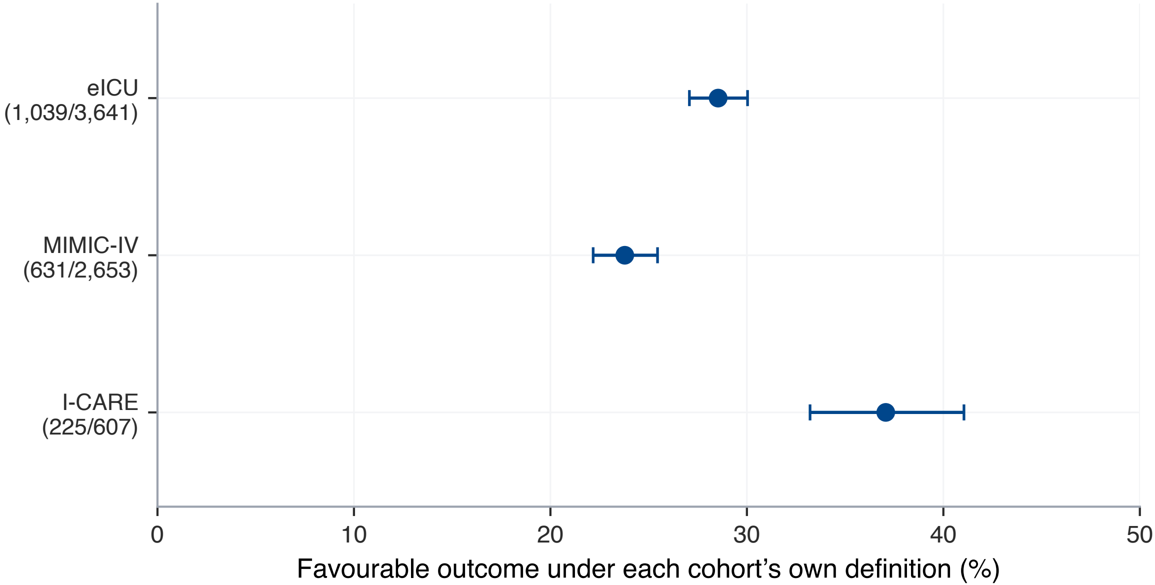
